# Spine Endoscopic Atlas-Tissue: multi-class anatomical segmentation in endoscopic spine surgery

**DOI:** 10.64898/2026.11.08.26367576

**Authors:** Yanhong Chen, Jianjin Zhang, Shan Wang, Qingxi Zhou, Xingyu Li, Jiapeng Li, Tianye Luo, Chaobo Feng, Zhouyang Hu, Jax Luo, Xiang Liao, Guoxin Fan, Mao Pang

## Abstract

Spine Endoscopic Atlas–Tissue (SEA-T) is a multicenter dataset for pixel-level multi-class semantic segmentation in endoscopic spine surgery. It contains 3,310 expert-annotated frames from 65 patients (1,663 cervical frames from 39 patients; 1,647 lumbar frames from 26 patients) across three centers, plus 11,155 unlabeled frames. Annotations cover nine classes—upper/lower lamina, ligamentum flavum, dural sac, nerve root, adipose tissue, muscle, disc material, and instrument channel—created in 3D Slicer under multi-stage quality control, with Fleiss’ kappa of 0.789. The annotated data are split by center, procedure, and anatomy into Center A training, internal validation, internal test, and held-out sub-cohorts, with Center B and Center C external tests; unlabeled frames are kept separate. Baselines spanning convolutional and foundation-model paradigms—nnU-Net v2, CaraNet, MedSAM2, and the SAM-based MA-SAM baseline—report Dice-based evaluation and reveal center-dependent domain shift. SEA-T is released with native-resolution PNG images, NRRD/MRML masks, RGB overlay images for annotation review, and metadata.

## Background & Summary

Spinal endoscopic surgery is a minimally invasive approach for disc discectomy, decompression, and fusion, and is widely used for degenerative cervical and lumbar diseases. Compared with open surgery, it can reduce postoperative pain and accelerate recovery, but the narrow field of view, low tissue contrast, bleeding, and instrument occlusion create a steep learning curve and increase the risk of anatomical misidentification[1]. These challenges make automated segmentation a key prerequisite for computer-assisted navigation, safety warning, and quantitative analysis of endoscopic spine procedures.

Recent progress in surgical AI has demonstrated the value of real-time anatomical recognition[2]. In spinal endoscopy, multi-class semantic segmentation of key cervical anatomical structures has been shown to be feasible in a single-center proof-of-concept study[3] [4], with CaraNet achieving a mean Dice of 0.865 on a held-out internal test set [5]. However, spinal endoscopy remains constrained by the scarcity of multicenter, expert-annotated, multi-class, pixel-level datasets [6] [7] [8]. Boundary annotation is costly, and spinal endoscopic images further exhibit center-dependent variation in endoscope optics, illumination, patient anatomy, and surgical workflow[9] [10]. Robust models therefore require diverse data with consistent anatomical definitions rather than larger numbers of weakly comparable labels.

Existing spinal endoscopic resources remain limited for anatomical semantic segmentation. Some datasets focus on instruments or mixed tissue-instrument instance annotations[11] [12], lack cervical coverage or multi-center stratification, and do not distinguish clinically relevant bony landmarks such as upper and lower lamina. Building on the Spine Endoscopic Atlas (SEA) [13] effort for instrument annotation in spinal endoscopy, we developed Spine Endoscopic Atlas–Tissue (SEA-T) as a complementary tissue annotation layer for multi-class semantic segmentation. SEA-T provides expert pixel-level masks for clinically relevant anatomical and operative-field classes, together with an unlabeled image pool for further semi-supervised or self-supervised learning [14]. The annotated cohort was collected across multiple centers using predefined patient-level internal and external splits to support evaluation of cross-center domain shift, with annotation consistency assessed by independent multi-rater labeling on a dedicated 200-frame reliability subset.

By releasing SEA-T, we aim to provide a reusable benchmark for anatomical structure segmentation in endoscopic spine surgery, enabling fair comparison of segmentation methods under realistic clinical variability. Detailed cohort construction, annotation protocol, quality control, split definitions, file formats, and baseline validation are described in Methods, Data Records, and Technical Validation.

## Methods

### Patient selection and clinical procedures

All surgical videos were retrospectively collected from 65 patients who underwent posterior endoscopic spine surgery for degenerative cervical or lumbar conditions at three referral centers between 2021 and 2024. The three contributing centers were Center A, Shenzhen Nanshan People’s Hospital, Shenzhen University Medical School, Shenzhen, Guangdong, China; Center B, Huidong Jianqiang Hospital, Huizhou, Guangdong, China; and Center C, The Third Affiliated Hospital of Sun Yat-sen University, Guangzhou, Guangdong, China. Center identifiers A/B/C are used consistently throughout the manuscript, metadata files, tables and figures. Procedures included percutaneous endoscopic interlaminar lumbar discectomy (PEID), unilateral biportal endoscopy (UBE), and posterior endoscopic cervical foraminotomy/discectomy (PECF).

The cohort comprised 1,663 cervical frames from 39 patients and 1,647 lumbar frames from 26 patients (total 3,310 annotated frames). Only primary single- or two-level cases in which the endoscope clearly visualized at least five of the nine target semantic classes for a cumulative duration of ≥3 minutes were included. Patients undergoing revision surgery, those with spinal deformity (scoliosis >20° or spondylolisthesis >Grade II), active infection, tumor, trauma, or severe intraoperative bleeding that obscured anatomy for >50% of the recording were excluded. This study was approved by the local ethics committees of the three participating centers (Center A: approval no. ky-2025-112901; Center B: approval no. HDJQK2026002; Center C: approval no. SL-II2026-352-02). Given the retrospective design and the use of strictly anonymized data, the requirement for informed consent was waived by these committees, which also approved the open publication of the de-identified dataset.

### Video acquisition and frame extraction

All procedures were recorded using standard endoscopic systems with native resolutions of 1280×720, 720×576, or 1920×1080 at 30 fps in H.264 format; a small subset of the Center C unlabeled pool comprises lower-resolution frames (352×288). Original recordings were exported without re-encoding. Frames were extracted at one frame per second using Python 3.12, ensuring sufficient temporal coverage while minimizing redundancy. After extraction, all frames were manually inspected. Images with severe motion blur, near-complete instrument occlusion (>50% of the field of view), or obvious loss of focus were excluded. Following this filtering process, the remaining eligible frames were imported into 3D Slicer, where the trained annotators performed pixel-level segmentation following the standardized protocol described below. This process yielded a total of 3,310 expertly annotated frames and 11,155 unlabeled frames. The complete data processing pipeline—from video recording to the final release-ready dataset—is summarized in Figure 1.

**Figure 1.**
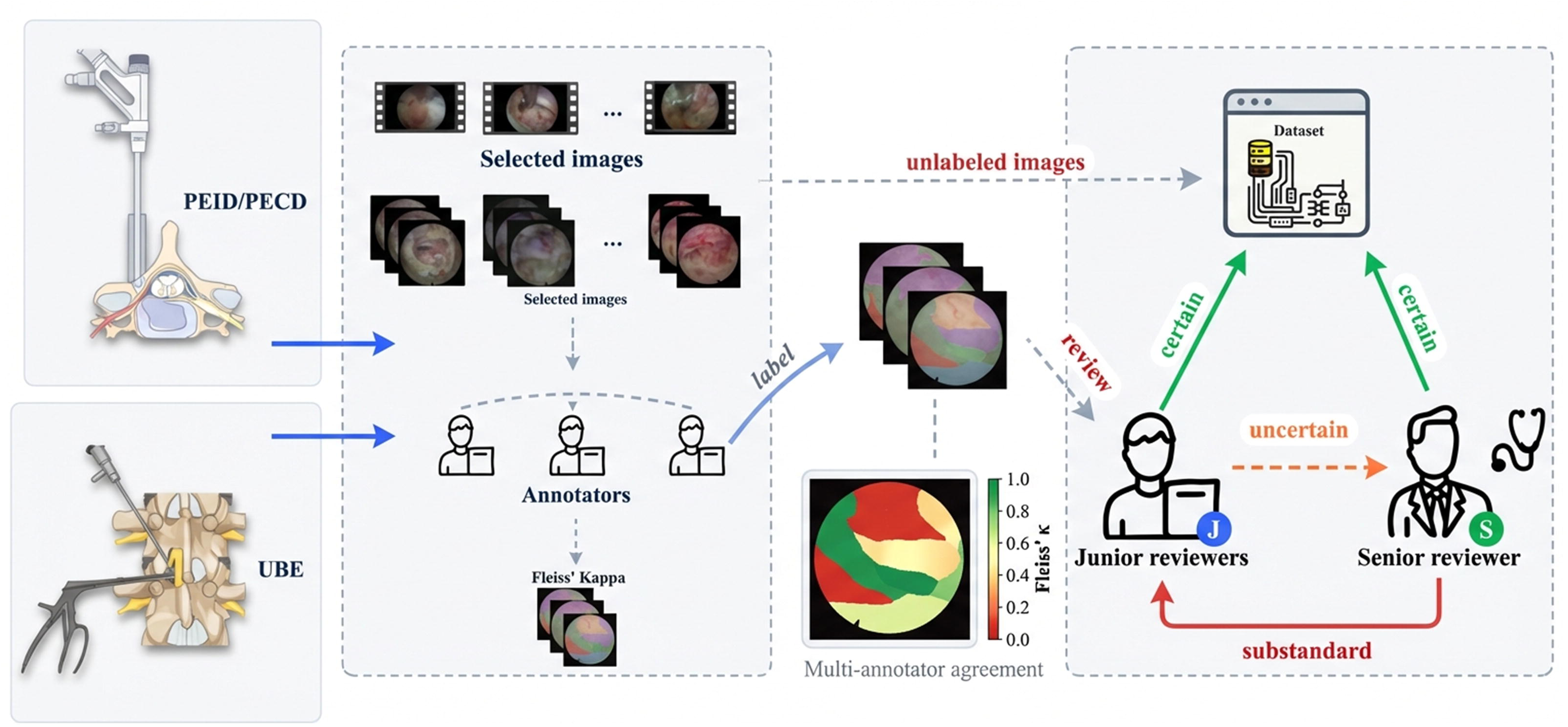
| Data curation and annotation workflow. Overview of the SEA-T dataset pipeline, from surgical video acquisition through frame extraction, quality filtering, multi-stage anonymization, pixel-level annotation in 3D Sheer, quality control, and final dataset partitioning into training, validation, internal test, external test, and unlabeled subsets.

### Image anonymization pipeline

To ensure compliance with applicable data protection regulations and institutional privacy policies, all frames underwent a multi-stage anonymization protocol. Video metadata were purged prior to extraction, and each frame was manually inspected by two independent reviewers to eliminate incidental reflections of personnel, text overlays, and identifying markers. The pipeline’s efficacy was validated on a random subset of 500 frames, confirming complete de-identification before dataset release.

### Annotation team and qualifications

Pixel-level annotation was performed by three trained annotators—full-time postgraduate researchers/doctors in spine and pain-related specialties—under the supervision of senior spine surgeons with extensive experience in posterior endoscopic spine surgery. To assess inter-rater agreement, 200 randomly selected frames were independently labeled by the three trained annotators; Fleiss’ kappa was calculated to evaluate the consistency of multi-rater annotations across all nine semantic classes (upper lamina, lower lamina, ligamentum flavum, dural sac, nerve root, adipose tissue, muscle, disc material, and channel).

### Annotation protocol and labeling hierarchy

The annotation process followed a standardized protocol to ensure consistency and accuracy in delineating the target semantic categories within the endoscopic field of view. Annotations were performed at the pixel level, assigning each pixel in the image to one of nine mutually exclusive semantic classes. These classes were selected based on their clinical relevance in posterior endoscopic spine surgery, focusing on structures commonly encountered during decompression and discectomy procedures. The labeling hierarchy prioritized foreground anatomical structures over background, with boundaries defined by visual cues such as texture, color gradients, and spatial relationships observed in the endoscopic images. Annotators were instructed to label only visible portions of structures, avoiding extrapolation into occluded areas, and to resolve ambiguities through reference to adjacent frames in the surgical video when necessary.

The classes and their corresponding labels used in the annotation software are:

- Ligamentum flavum (ligament): yellowish fibrous ligament between laminae; includes hypertrophic portions.
- Nerve root (nerve_root): tubular whitish structure with longitudinal striations; labeled along its sheath.
- Dural sac (dura): bluish-white, smooth, pulsating membrane; outline follows visible contour.
- Upper lamina (upper_lamina): superior bony edge of the vertebral arch; dense whitish structure.
- Lower lamina (lower_lamina): inferior bony edge of the vertebral arch; distinguished by position relative to the interlaminar window.
- Muscle (muscle): reddish-brown paraspinal fibers with striations
- Adipose tissue (adipofascia): loose yellowish fat with lobulated appearance; excludes vessels.
- Disc material (disc): whitish, gelatinous extruded or exposed disc fragments.
- Instrument (channel): instruments, blood, irrigation fluid, smoke, or unidentifiable regions.
- Although the channel class does not represent a specific anatomical structure, it was intentionally defined as a separate semantic category because the operative channel is frequently present alongside anatomical structures during spinal endoscopic procedures. Even after excluding identifiable surgical instruments, this region may contain transient contents such as irrigation fluid, blood, smoke, or visually ambiguous areas associated with the working pathway. Treating the channel as an independent class prevents these non-anatomical regions from being incorrectly assigned to surrounding anatomical structures and enables more accurate pixel-level semantic representation of the endoscopic surgical field.

Boundary rules emphasized sub-pixel precision, using magnification tools during annotation to trace edges along natural gradients. In cases of partial occlusion (e.g., by blood or instruments covering <50% of a structure), annotators labeled the visible segments and marked transitions as “soft” boundaries. Representative examples of the nine semantic classes included in SEA-T are shown in Figure 2a, illustrating both whole-frame annotations and magnified regions highlighting boundary delineation. Overlapping structures were resolved by hierarchical priority (Figure 2b): neural elements (dural sac, nerve root) take precedence over bony/ligamentous tissues (ligamentum flavum, upper/lower lamina, disc), which in turn supersede soft tissues (muscle, adipose tissue). Instrument channel occupies the lowest priority as the default category for instruments, fluids, and non-anatomical regions. The channel class was introduced to represent non-anatomical operative regions frequently encountered during endoscopic procedures and to reduce false-positive anatomical predictions.

**Figure 2.**
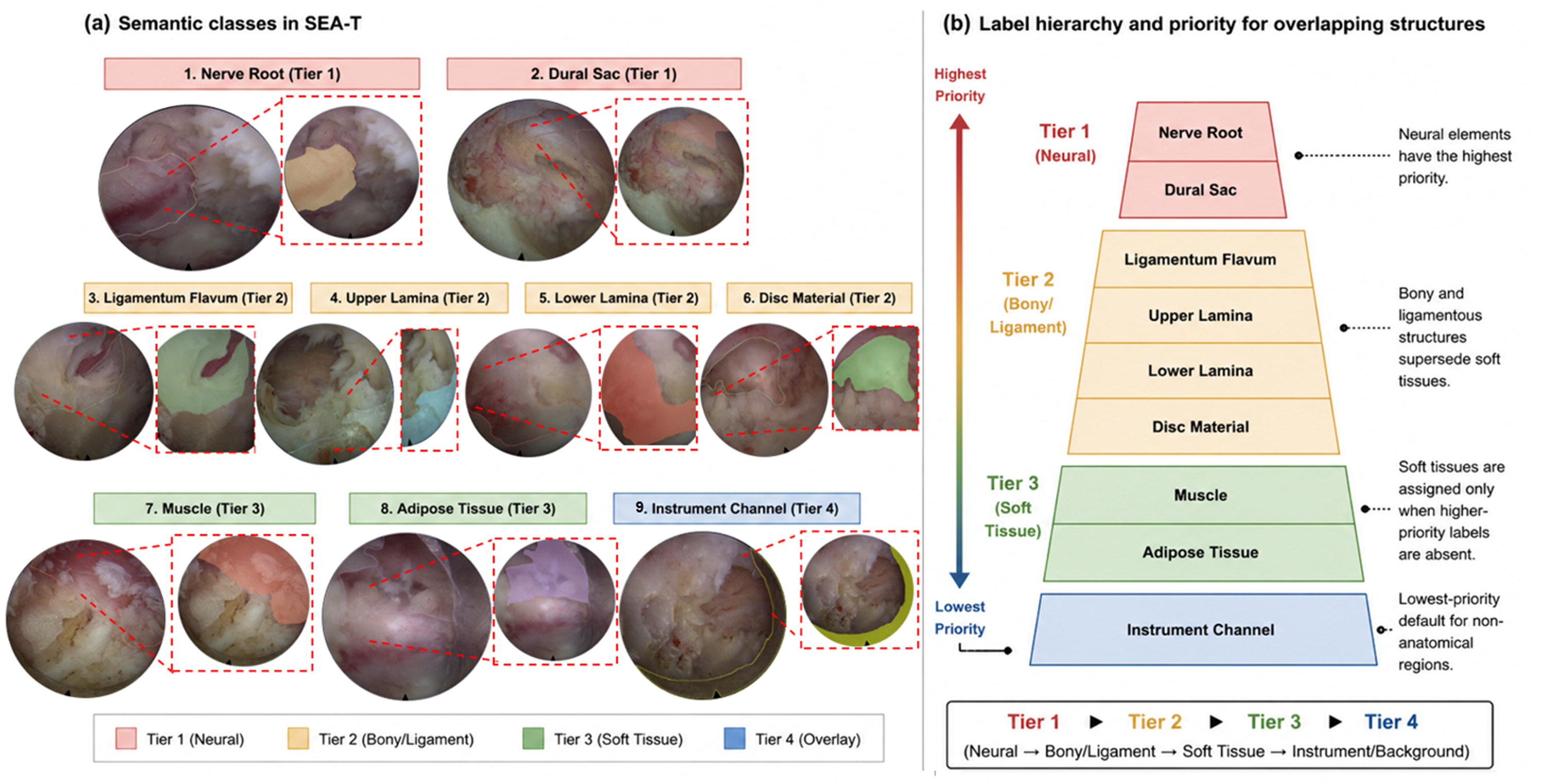
| Semantic classes and labeling hierarchy, (a) Representative examples of the nine semantic categories in SEA-T, showing whole-frame annotations and magnified insets highlighting boundary delineation for upper lamina, lower lamina, ligamentum flavum, dural sac, nerve root, adipose tissue, muscle, disc material, and instrument channel, (b) Hierarchical priority scheme for resolving overlapping structures: neural elements take precedence over bony and ligamentous tissues, which in turn supersede soft tissues, with instrument channel assigned lowest priority.

### Annotation tools and quality control workflow

All anatomical annotations were performed using 3D Slicer (version 5.6.1), chosen for its robust support for pixel-level manual segmentation, flexible module-based workflow, and compatibility with high-resolution endoscopic imaging. Frames were imported in their native resolutions (1,280 × 720, 720 × 576, or 1,920 × 1,080), and annotators used the Segment Editor module to generate structure-specific masks. Polygon-based drawing, paint, and erase tools were applied iteratively to delineate boundaries, preserving anatomical fidelity. This environment enabled consistent layer-wise segmentation for curved structures such as ligamentum flavum and dural sac, where precise contouring is essential to downstream model development.

A structured, multi-stage QC workflow was implemented to maintain consistency across annotators. First, each batch of 50 frames was screened within 3D Slicer using synchronized comparison views, and custom Python scripts were used to flag recurrent issues such as discontinuous boundaries, topological violations, fragmented nerve root trajectories, or isolated label islands. Annotators then performed reciprocal review of one another’s masks; disagreements—typically involving boundary placement in low-contrast regions or interpretation of partially obscured tissue—were discussed collaboratively, and frames without consensus were submitted to a senior spine surgeon for adjudication. Finally, a senior surgeon performed an independent, blinded audit of 20% of each batch; issues identified during audit were addressed in regular QC meetings to standardize annotation criteria and reduce inter-observer variability. To mitigate fatigue-related error, annotation sessions were limited to two hours with mandatory breaks, and workload was balanced across annotators.

Before final inclusion, all masks were exported as NRRD label maps and as RGB overlay images, which were visually compared against the corresponding frames. This final inspection focused on detection of editing artifacts—such as aliasing, residual brush patterns, or inadvertent smoothing—ensuring that each segmentation met the predefined anatomical and technical quality standards required for downstream modeling and multi-center validation. The details of the entire workflow are shown in Figure 1.

### Inter-rater reliability assessment

Inter-rater reliability was evaluated using Fleiss’ kappa coefficient, a statistical measure chosen because multiple annotators independently assigned categorical labels to each pixel; Dice similarity coefficient, while standard for segmentation accuracy evaluation, is not designed for measuring inter-observer agreement among independent raters [15]. Fleiss’ κ quantifies agreement beyond chance across the nine semantic classes and is suitable for multi-rater categorical data. A subset of 200 frames (randomly selected from the 3,310 annotated frames) was independently labeled by the three trained annotators, without access to prior labels or discussions.

Fleiss’ kappa was calculated [16] [17] for each class using the formula:

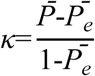

where *P̅* is the average observed agreement, and *P̅_e_* is the average agreement expected by chance. Computations were performed in Python using the statsmodels package. Agreement was substantial to almost perfect across classes (overall κ = 0.789, P < 0.001); class-wise values and analyses are reported in Technical Validation, Figure 3, and Table 1. After the reliability assessment, discordant regions (∼5% of pixels) were re-annotated through group consensus before finalizing the dataset. Only the consensus-refined annotations are released; multi-rater masks from the reliability subset are not included in the current release.

**Figure 3.**
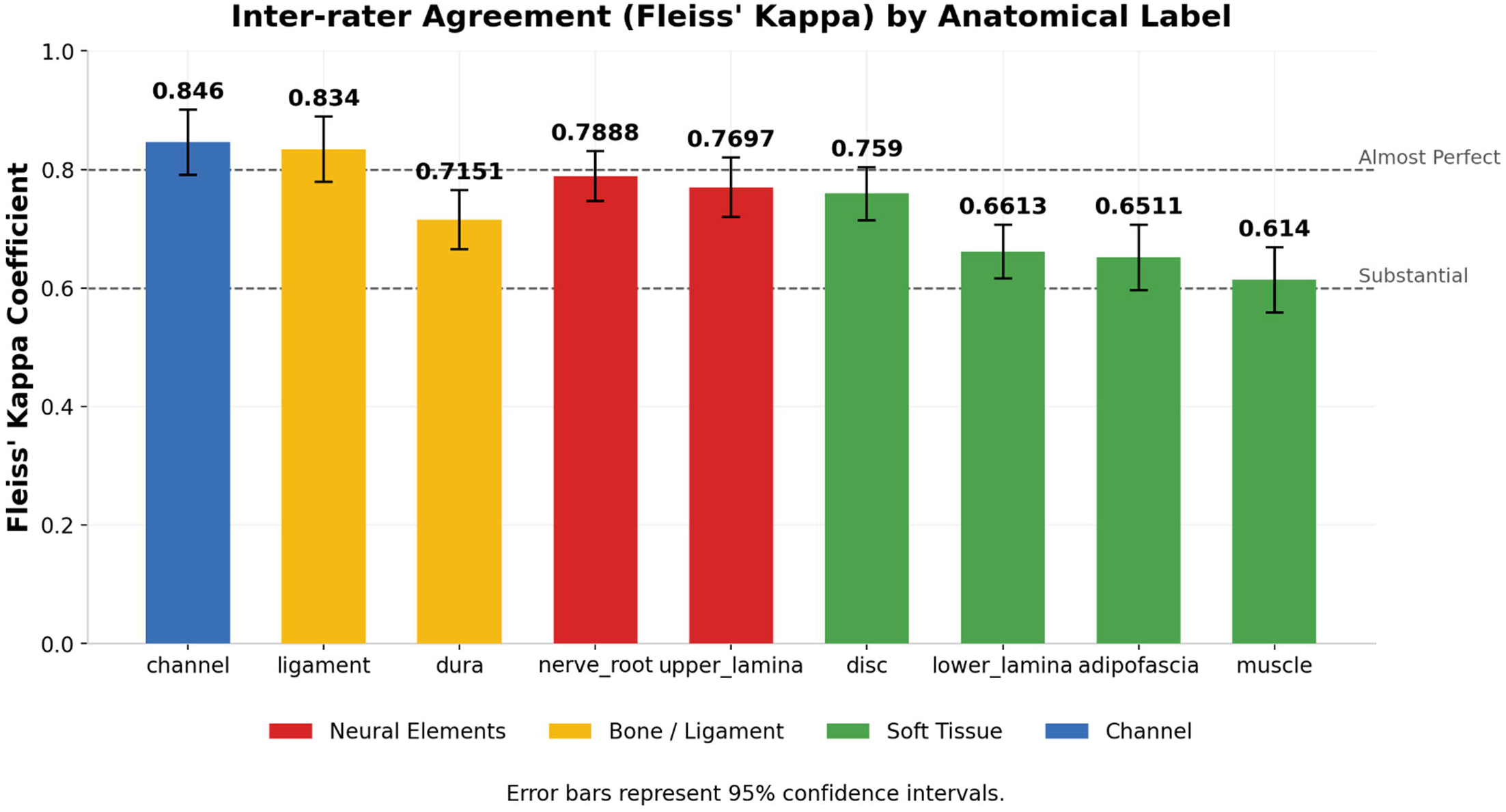
| Inter-rater reliability across semantic classes. Fleiss’ kappa coefficients for pixed-level agreement among three independent annotators (n = 20 frames), with class-specific values ranging from 0.614 (muscle) to 0.846 (instrument channel); overall mean k = 0.789 (P < 0.001). Error bars denote 95% confidence intervals.

**Table 1.**
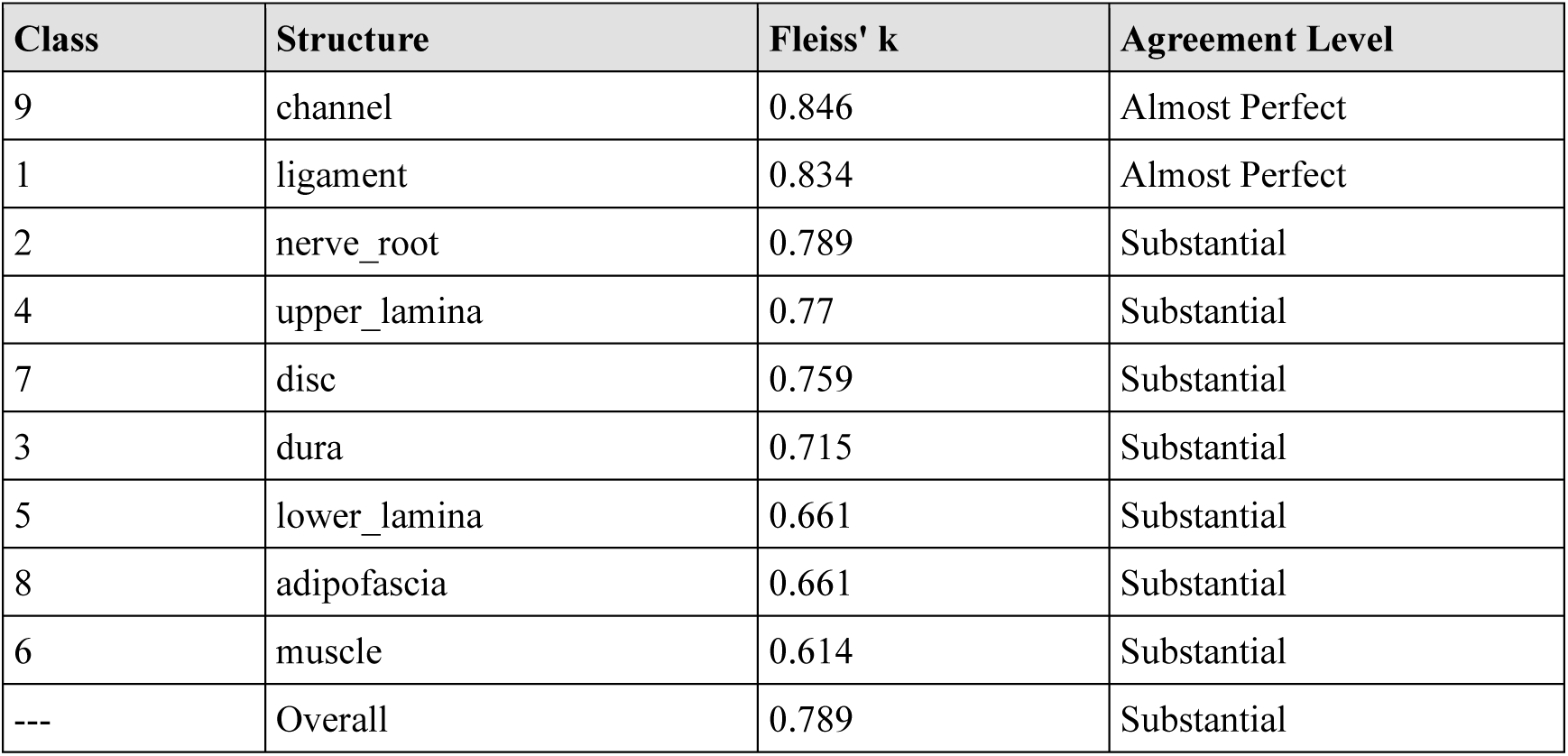
Inter-rater reliability assessment for pixel-level anatomical segmentation in SEA-T. Fleiss’ Kappa with agreement levels: Substantial (0.61-0.80), Almost Perfect (0.81-1.00)

| **Class** | **Structure** | **Fleiss' k** | **Agreement Level** |
|---|---|---|---|
| 9 | channel | 0.846 | Almost Perfect |
| 1 | ligament | 0.834 | Almost Perfect |
| 2 | nerve_root | 0.789 | Substantial |
| 4 | upper_lamina | 0.77 | Substantial |
| 7 | disc | 0.759 | Substantial |
| 3 | dura | 0.715 | Substantial |
| 5 | lower_lamina | 0.661 | Substantial |
| 8 | adipofascia | 0.661 | Substantial |
| 6 | muscle | 0.614 | Substantial |
| --- | Overall | 0.789 | Substantial |

### Dataset splitting strategy

For the baseline experiments in this study, the 3,310 annotated frames were divided following a stratified approach by center, procedure type (PECF/PEID/UBE), and spinal region, enabling reproducible training, internal evaluation, and rigorous assessment of cross-center generalization [10]. The released dataset itself does not impose a fixed partition; the per-frame metadata file allows users to define their own splits as needed. The partition adopted for the baselines is described below.:

#### 1) Internal cohort

Center A: 2,586 annotated frames from 50 patients; 429 frames (12 patients, “without training” subset) were additionally annotated after the initial training phase and are released as an independent held-out sub-cohort that was never involved in model development; the remaining 2,157 frames (38 patients) were split as follows:

- Training set: 70% (1,510 frames from 26 patients).
- Internal validation set: 15% (324 frames from 6 patients).
- Internal test set: remaining 15% (323 frames from 6 patients).

#### 2) External cohort Center B

- Cervical external test set: 200 frames (6 patients, PECF).
- Lumbar external test set: 303 frames (7 patients, PEID).

Center C:

- Lumbar external test set: 221 frames (2 patients, UBE).

All annotated frames from Center B (503 frames) and Center C (221 frames) were reserved exclusively as an external test set. This subset was never seen during model development or hyperparameter tuning and serves to quantify domain shift caused by differences in endoscope optics, illumination, surgical technique, and patient population.

The large pool of unlabeled frames (11,155 images from Centers A and C) was kept entirely separate and is provided without masks for use in further semi-supervised, self-supervised, or weakly supervised experiments.

### Data storage format and directory structure

The SEA-T dataset is released in a lossless, open format that retains the original endoscopic appearance from three centers and can be loaded directly by any standard framework.

Images are provided in their native resolutions without resizing or color modification:

- Center A: 1280 × 720 RGB PNG (8-bit per channel)
- Center B: 720 × 576 RGB PNG (8-bit per channel) for the lumbar subset; the 200 cervical external test frames are provided at 1280 × 720
- Center C: 1280 × 720 or 1920 × 1080 RGB PNG (8-bit per channel)

For 3,310 annotated frames, segmentation masks were generated in 3D Slicer and archived as NRRD files; companion MRML scene files retain annotation metadata. For visual reference, RGB PNG overlays are additionally exported at identical resolution to source images, in which annotated anatomical and operative-field boundaries are rendered as colored contours with colors assigned per class as defined in Table 2, preserving annotation order and nomenclature.

**Table 2.** Class label encoding for pixel-level semantic segmentation in SEA-T. Label IDs and RGB colors extracted from NRRD segmentation files (3D Slicer default color coding)

| **Label ID** | **Structure** | **R** | **G** | **B** | **Hex Color** | **Color** |
|---|---|---|---|---|---|---|
| 0 | background | — | — | — | — | — |
| 1 | ligament | 128 | 174 | 128 | #80AE80 |  |
| 2 | nerve root | 241 | 214 | 145 | #F1D691 |  |
| 3 | dura | 177 | 122 | 101 | #B17A65 |  |
| 4 | upper lamina | 111 | 184 | 210 | #6FB8D2 |  |
| 5 | lower lamina | 216 | 101 | 79 | #D8654F |  |
| 6 | muscle | 221 | 130 | 101 | #DD8265 |  |
| 7 | disc | 144 | 238 | 144 | #90EE90 |  |
| 8 | adipofascia | 183 | 156 | 220 | #B79CDC |  |
| 9 | channel | 220 | 245 | 20 | #DCF514 |  |

For baseline evaluation in this study, the annotated frames were partitioned into training, validation, internal testing, and external testing sets, as detailed in the Dataset splitting strategy. The released dataset does not impose a fixed partition; annotated and unlabeled frames are stored in separate directories, allowing users to design their own splits. A structured per-frame metadata file provides information, including center origin, patient identifier, spatial resolution, procedure type, spinal level, and annotation availability, allowing users to perform fine-grained filtering and stratified sampling. Figure 4 presents the full directory structure of SEA-T, reflecting the organization described above.

**Figure 4.**
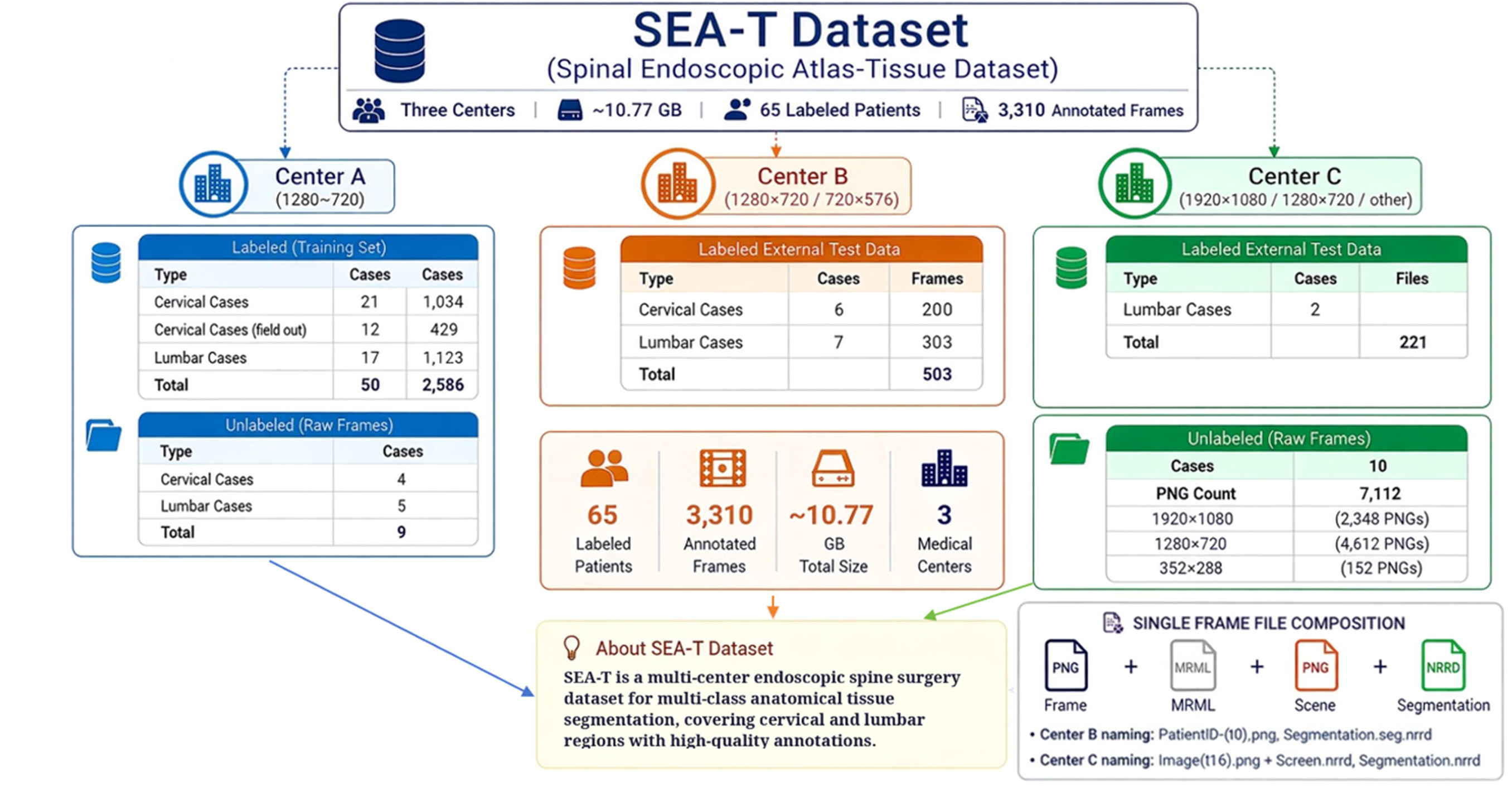
| SEA-T dataset overview and organization. Distribution of labeled and unlabeled frames across three centers: Center A (labeled subsets, held-out subsets and unlabeled subsets), Center B (cervical and lumbar external test), and Center C (lumbar external test and unlabeled pool). Each annotated frame comprises a native-resolution RGB image (PNG), thumbnail (PNG), 3D Sheer scene file (MRML), and segmentation mask (NRRD). The full dataset comprises 3,310 annotated frames from 65 patients and 11,155 unlabeled frames, totaling approximately 10.77 GB.

This structure, together with the patient-level metadata, facilitates leakage-free split design, clearly indicates resolution differences, and allows immediate use with standard medical segmentation frameworks such as nnU-Net v2[18], CaraNet[19], MedSAM2 [20] [21] and MA-SAM [22].

The complete dataset occupies approximately 10.77 GB when extracted.

### Data Records

#### 1. Annotated Image-Mask Pairs

This record contains 3,310 expertly annotated frames with pixel-level segmentation masks across nine semantic classes, derived from 65 patients. Files are organized as: Images: Native resolution RGB frames in PNG format

- Cervical: 1,663 frames (1280×720)
- Lumbar: 1,647 frames (1280×720, 720×576, or 1920×1080)
- Masks/NRRD: 3D Slicer-compatible volumetric masks (.nrrd)
- MRML_scenes: 3D Slicer scene files for annotation verification

Native resolutions vary by center: Center A (1280×720); Center B (1280×720 for cervical and 720×576 for lumbar external test frames); and Center C (1280×720 or 1920×1080). All masks are provided as NRRD label maps at identical resolution to source images, with voxel values encoding class labels according to Table 2.

#### 2. Unlabeled Image Pool

A collection of 11,155 frames passing quality control but lacking annotations, provided in PNG format for semi-supervised or self-supervised learning research.

#### 3. Metadata

A structured per-frame metadata file (metadata.csv) accompanies the dataset, with one row per frame and the following fields: a unique frame identifier; contributing center; clinical category; anonymized patient identifier; relative paths to the native-resolution RGB image, the 3D Slicer scene files (scene image and companion MRML scene), and the NRRD segmentation mask; image resolution; and per-file sizes (image, mask, and total). Patients are referenced only by anonymized codes (e.g., P1, P2), and no directly identifying information is included. The directory hierarchy separates centers, annotation status, and mask–image pairs to prevent accidental data leakage (Figure 4).

Class distribution across the nine semantic categories is summarized in Table 3, reflecting surgical phase and field-of-view dynamics.

**Table 3.** Class distribution in the SEA-T benchmark splits. Proportions are relative to foreground classes 1–9 only; background is excluded. n=2,881 annotated frames across the benchmark splits (Center A: 2,157; Center B: 503; Center C: 221), excluding the 429-frame Center A held-out sub-cohort.

| **Label ID** | **Structure** | **Frame Count** | **Frequency (%)** | **Total Pixels** | **Avg Pixels / Frame** | **Proportion (%)** |
|---|---|---|---|---|---|---|
| 1 | ligament | 2,457 | 85.3 | 321,952,439 | 131,035 | 30.69 |
| 5 | lower lamina | 1,974 | 68.5 | 173,849,095 | 88,069 | 16.57 |
| 4 | upper lamina | 1,805 | 62.7 | 157,961,830 | 87,513 | 15.06 |
| 2 | nerve root | 1,425 | 49.5 | 88,020,102 | 61,768 | 8.39 |
| 3 | dura | 1,321 | 45.9 | 97,306,213 | 73,661 | 9.28 |
| 7 | disc | 998 | 34.6 | 80,209,201 | 80,370 | 7.65 |
| 8 | adipofascia | 935 | 32.5 | 85,885,069 | 91,856 | 8.19 |
| 6 | muscle | 497 | 17.3 | 32,944,977 | 66,288 | 3.14 |
| 9 | channel | 295 | 10.2 | 10,763,349 | 36,486 | 1.03 |
| — | Total foreground | 2,881 | 100.0 | 1,048,892,275 | 364,072 | 100.00 |

*Note: Foreground = classes 1–9 (ligament through channel). Background (label 0) is not manually annotated and is excluded from proportions.*

### Technical Validation

#### Annotation quality and inter-rater reliability

Annotation quality was ensured through a multi-stage quality-control workflow in which pixel-level segmentation was performed by three trained annotators under the supervision of senior spine surgeons, with reciprocal peer review, expert adjudication of unresolved disagreements, and blinded audit of 20% of each batch applied throughout (Methods). Inter-rater reliability, assessed on 200 frames independently labeled by all three annotators, yielded a Fleiss’ κ of 0.789 (P < 0.001) across the nine semantic classes, with class-wise values ranging from 0.614 (muscle) to 0.846 (instrument channel) (Table 1). The lower agreement for muscle reflects its variable texture and limited occurrence, whereas bony structures and instruments achieved the highest consistency. Discordant regions (affecting ∼5% of pixels) were subsequently re-annotated through group consensus to refine the final dataset. These results indicate that SEA-T provides reproducible pixel-level annotations suitable for supervised learning and quantitative benchmarking.

#### Dataset diversity and clinical coverage

The dataset spans three independent centers using different endoscopic systems and acquisition settings, covering both cervical (PECF) and lumbar (PEID/UBE) procedures across 65 patients (native resolutions are summarized in Data Records). All nine semantic classes are represented across multiple patients and procedures, and the surgical videos capture clinically realistic variability—including bleeding, illumination variation, partial instrument occlusion, and low-contrast tissue interfaces—that poses substantial challenges for automated segmentation. Importantly, all centers followed the identical annotation protocol and labeling scheme, ensuring that observed performance differences reflect true acquisition-related variation rather than inconsistent ground-truth standards. This heterogeneity, preserved rather than harmonized away, provides the diversity needed to evaluate model robustness under realistic clinical conditions.

#### Baseline segmentation experiments

To assess dataset compatibility rather than to introduce a new architecture, we evaluated four publicly available segmentation frameworks spanning convolutional and foundation-model paradigms: nnU-Net v2, CaraNet, MedSAM2 and MA-SAM. All models were trained using their original recommended configurations on the 70/15/15 Center A split. Because the released evaluation pipelines of these frameworks report results at different granularities, evaluation follows two conventions (Table 4): MA-SAM and nnU-Net were assessed at the case level using GT-present mean Dice[15], whereas MedSAM2 and CaraNet were assessed using the class-wise one-vs-rest slice Dice of their released pipelines, which do not implement case-volume aggregation; unifying all four frameworks under case-volume aggregation is planned for a future benchmark revision. Accordingly, the results should be compared within the same metric family rather than directly across the two conventions—for example, the apparently higher MedSAM2 scores in external Center B lumbar cases reflect slice-level rather than case-level aggregation and should not be read as superior case-level performance. The baselines converged stably without class collapse and produced anatomically meaningful predictions, indicating that SEA-T is compatible with representative segmentation tools. Class-wise analysis of the GT-present nnU-Net baseline supported a clinically realistic difficulty gradient: ligament, adipofascia and channel were generally more reproducible, whereas nerve root was consistently among the most difficult classes. Class rankings remained center- and anatomy-dependent—for example, channel performance dropped markedly in external Center B lumbar cases—highlighting the importance of reporting class-wise and center-wise metrics rather than relying on a single global Dice[15]. The 429-frame held-out sub-cohort (12 patients) was reserved for future benchmark iterations and was not used in the present baseline evaluation.

**Table 4.**
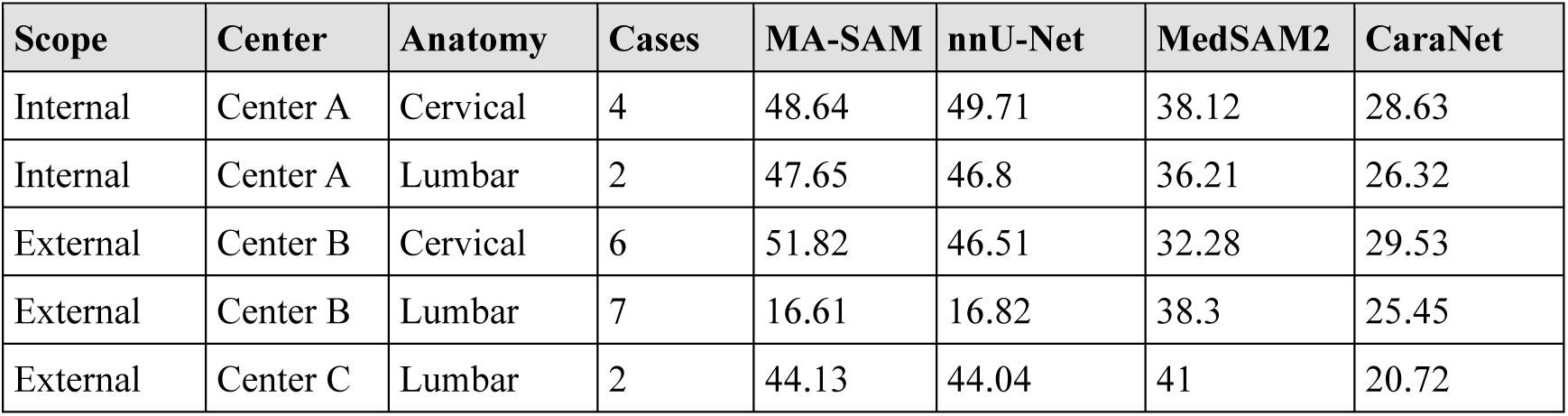
Baseline segmentation performance (GT-present case-volume Dice, %) on the internal and external test subsets. MA-SAM and nnU-Net use GT-present case-volume Dice; MedSAM2 and CaraNet use class-wise one-vs-rest slice Dice.

| **Scope** | **Center** | **Anatomy** | **Cases** | **MA-SAM** | **nnU-Net** | **MedSAM2** | **CaraNet** |
|---|---|---|---|---|---|---|---|
| Internal | Center A | Cervical | 4 | 48.64 | 49.71 | 38.12 | 28.63 |
| Internal | Center A | Lumbar | 2 | 47.65 | 46.8 | 36.21 | 26.32 |
| External | Center B | Cervical | 6 | 51.82 | 46.51 | 32.28 | 29.53 |
| External | Center B | Lumbar | 7 | 16.61 | 16.82 | 38.3 | 25.45 |
| External | Center C | Lumbar | 2 | 44.13 | 44.04 | 41 | 20.72 |

#### Domain shift assessment

External evaluation on the held-out test cohorts revealed substantial center-dependent variability (Table 4). Mean Dice scores dropped markedly on Center B (720 × 576, external lumbar, 7 patients), whereas Center C (1,280 × 720/1,920 × 1,080), which matches or exceeds the internal cohort’s native resolution, yielded considerably higher scores. This dichotomy indicates that resolution-mediated acquisition shift constitutes a dominant factor in cross-center performance[23]. These results demonstrate that SEA-T preserves realistic acquisition variability and enables evaluation of model robustness under cross-center domain shifts.

#### Representative segmentation examples

Qualitative inspection of model predictions (Figure 5) illustrates that SEA-T contains both straightforward and challenging anatomical scenarios, including blurred tissue interfaces, partial instrument occlusion, illumination variation, and low-contrast neural structures. Corresponding quantitative results (Table 4) show GT-present case-volume mean Dice of 49.71% (cervical) and 46.80% (lumbar) for nnU-Net and 48.64% and 47.65% for MA-SAM on the internal test set, while structure-specific performance ranged from 83–85% for ligamentum flavum and instrument channel to ≤31.72% for nerve root. These results demonstrate that SEA-T preserves realistic anatomical difficulty while supporting quantitative benchmarking.

**Figure 5.**
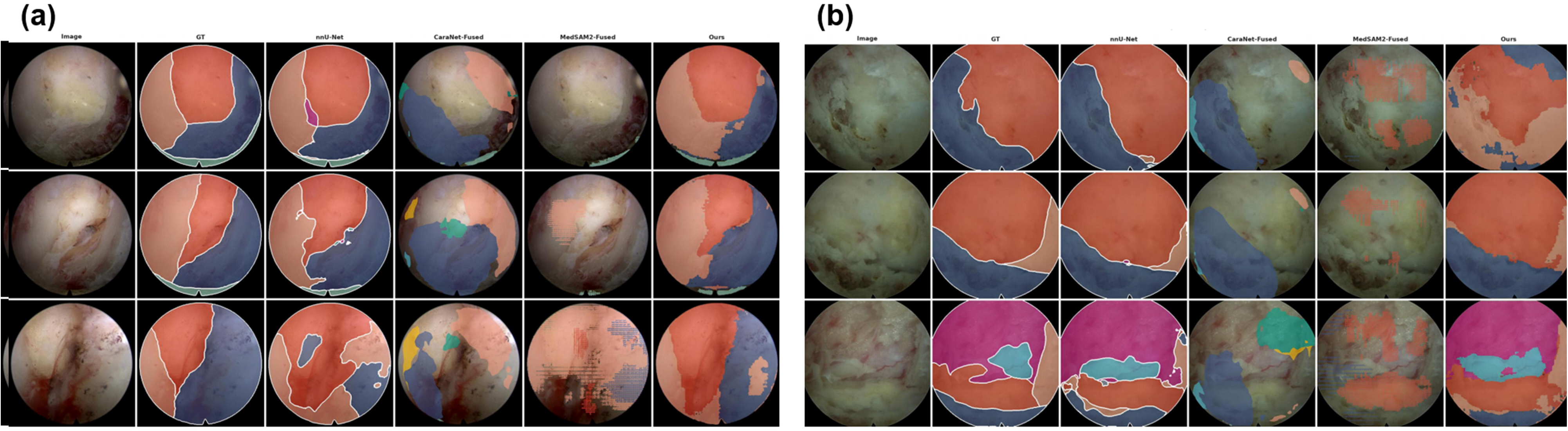
| Representative segmentation predictions on the internal test set. (a) Cervical and (b) lumbar cases showing qualitative comparison of ground-truth masks and model predictions (nnU- Net and MA-SAM), illustrating typical scenarios including clear tissue interfaces, partial instrument occlusion, illumination variation, and low-contrast neural structures.

#### Reproducibility and data accessibility

All data are released in lossless, open formats as described in Data Records, together with the frame-level baseline split definitions and the frame extraction and pre-processing scripts used in this study; all benchmarked methods (nnU-Net, CaraNet, MedSAM2, and MA-SAM) were used via their original, publicly available implementations, with no custom model code.

### Usage Notes

#### Recommended applications

SEA-T was designed primarily for research on computer-assisted spinal endoscopic image analysis, including semantic segmentation, anatomical structure recognition, surgical scene understanding, and domain generalization. The dataset supports both fully supervised learning using expert-annotated image–mask pairs and semi-supervised or self-supervised learning using the additional unlabeled image pool.

The annotated subset can be used for developing and benchmarking pixel-level segmentation algorithms targeting nine clinically relevant semantic classes: upper lamina, lower lamina, ligamentum flavum, dural sac, nerve root, adipose tissue, muscle, disc material, and operative field/non-anatomical regions. The unlabeled image pool provides opportunities for representation learning, foundation model adaptation, and semi-supervised segmentation approaches.

#### Dataset organization and loading

The dataset is distributed in a lossless format preserving native endoscopic image characteristics. RGB images are provided as PNG files, while segmentation masks are available as both 3D Slicer-compatible NRRD files and RGB overlay images with class-coded contours and MRML scene files are additionally provided. The accompanying structured per-frame metadata provides frame-level information, including center origin, surgical procedure type, spinal region, image resolution, and annotation availability.

Researchers should use the predefined patient-level split for benchmarking to prevent information leakage caused by highly correlated frames extracted from the same surgical videos. The external test cohorts from independent centers should remain reserved for evaluating cross-center generalization.

#### Recommended preprocessing and training strategies

SEA-T intentionally preserves acquisition heterogeneity across centers, including differences in endoscope systems, image resolution, illumination conditions, and surgical workflows. Therefore, users are encouraged to report preprocessing procedures explicitly and avoid aggressive normalization strategies that may eliminate clinically relevant domain variation.

The dataset is compatible with commonly used medical segmentation frameworks, including nnU-Net, convolutional neural network-based methods, and foundation model-based approaches. Models should be evaluated using class-wise metrics in addition to overall segmentation performance, as anatomical structures in spinal endoscopy exhibit substantial differences in size, appearance, and segmentation difficulty.

#### Evaluation considerations

Due to the dynamic nature of spinal endoscopic imaging, certain anatomical structures, particularly nerve roots and partially occluded tissues, represent challenging segmentation targets. Performance comparisons should consider class imbalance, anatomical visibility, and center-specific acquisition characteristics rather than relying solely on global Dice similarity coefficients [15].

For fair comparison, we recommend reporting:

1. overall segmentation performance.
2. class-specific Dice scores.
3. performance across cervical and lumbar procedures.
4. internal and external center evaluation results.
5. qualitative visualization of anatomically critical structures.

## Data availability

The dataset, annotation files, and metadata have been deposited on figshare (DOI: https://doi.org/10.6084/m9.figshare.34767681) and will be released under the Creative Commons Attribution 4.0 International license (CC BY 4.0). The record is currently under embargo and will be made publicly available upon publication. Frame extraction and pre-processing scripts (Python 3.12; OpenCV and NumPy) are provided together with the dataset files.

## Code availability

All baseline experiments in this study were performed using the original, publicly available implementations of the respective methods, with no custom model code:

nnU-Net: https://github.com/mic-dkfz/nnunet

CaraNet: https://github.com/AngeLouCN/CaraNet

MedSAM2: https://github.com/bowang-lab/MedSAM2

MA-SAM: https://github.com/cchen-cc/MA-SAM.

## Author Contributions

Y.C., J.Z., S.W., and Q.Z. contributed equally to this work. Conceptualization: X.L., G.F., and M.P. Methodology: Y.C., J.Z., S.W., Q.Z., Jx.L., and M.P. Data curation: Y.C., J.Z., J.L., C.F., S.W., and Q.Z. Software and formal analysis: J.Z., Z.H., and Jx.L. Validation: Y.C., Z.H., S.W., Jx.L., and Q.Z. Investigation and resources: T.L., C.F., J.L., S.W., Q.Z., M.P., G.F., and X.L. Project administration: Y.C., J.L., and Q.Z. Visualization: J.Z. and Y.C. Supervision: Jx.L., M.P., X.L., and G.F. Funding acquisition: M.P., X.L., and G.F. Writing—original draft: Y.C. and J.Z. Writing—review and editing: all authors. All authors read and approved the final manuscript.

## Acknowledgements

We thank the surgeons, clinical teams, and annotators at the participating centers for their contributions to surgical video collection, anonymization, annotation, and quality control. We also thank the patients whose de-identified data made this resource possible.

